# Do MOXFQ scores change over time? A retrospective study of podiatric surgery outcomes

**DOI:** 10.1101/2023.08.10.23293952

**Authors:** Victoria North, Ian Reilly, Andy Bridgen, Anthony Maher

## Abstract

**Background:** Recording patient-reported outcome measurements six months after surgery using PASCOM-10 is common practice in podiatric surgery in the United Kingdom. This study aims to establish if the health-related quality of life after foot surgery is changed at the 12- month point.

**Method:** An audit of patient-reported outcomes following foot surgery was undertaken. Electronic case notes were examined, and 236 patients were identified over 1 year. Patients completed the Manchester Oxford Foot/Ankle Questionnaire (MOXFQ) pre-surgery, at six months post-operatively (in person/telephone conversation) and were then invited by letter to complete this again at 12 months post-operatively.

**Results:** 91 participants (39%) completed both post-operative questionnaires (66 females 25 males), 45% completed a six-month questionnaire only and 61% completed a 12-month questionnaire. The results demonstrate a significant score change between the three time points and across the three MOXFQ domains of walking/standing, pain and social interaction (0.353, F [6, 356] = 40.59, p <.001), and a further interaction with gender (0.435, F [6,352] = 30.23, p <.001). An average, positive change in health-related quality of life (HRQOL) across all domains was the greatest from pre-surgery to six months post-surgery at 78% improved, and then decreased to 62% improved at 12 months post-surgery. Females experience lower HRQOL before surgery, at 6 months and at 12-month post-operation time points but gain the greatest improvement over pre-operative scores at 30%.

**Conclusion:** A patient-centred approach is key in foot surgery, patient-reported health-related quality of life changes significantly at different time points. In this review, the overall health-related quality of life improved with surgery at the six- and 12-month point, but with some deterioration at 12 months. A short period of recovery may not yield genuine long-term patient-reported outcomes, and an increase to a minimum of 12 months is recommended from the more common six months. However, the different methods of collecting post-operative data at various time points may have had an influence on scores.

**Level of clinical evidence**:4

## Introduction

Foot and ankle pathology account for a significant percentage of all orthopaedic interventions. Podiatric surgery is concerned with the surgical management of foot and ankle conditions and has grown in scope of practice since the encouragement of day case surgery in the 1990s (1), with day surgery saving the NHS around £2 billion from 1998 to 2013 (2).

The NHS takes a patient-centred approach to health care with an estimated 1.5 million people being supported daily by health services in the UK (3). There are numerous health outcome tools available for use in foot surgery. Patient-reported outcomes measures (PROMs) that are appropriately designed and condition-specific, which are reliable and valid, focus on the health concerns of most relevance to patients, such as pain, function, and mobility. Collectively these factors form a patient’s health-related quality of life (HRQOL) (4,5). The tools must be responsive to change, reproducible and reflective of the patient’s perception (6), and some authors suggest a shift toward the consistent use of a smaller number of valid, reliable, and clinically valuable scales (7).

The MOXFQ is the preferred standard instrument of the Royal College of Podiatry (8), utilised through the Podiatric Audit of Surgery and Clinical Outcome Measurement (PASCOM-10) system which the college recommends for all Podiatric Surgery teaching units in the UK (6). The MOXFQ is completed pre-surgery and six months post-operative as standard (9) but there are no formal guidelines or best practice available on recommended time intervals, with limited studies evaluating time points as a primary indicator for outcomes. However, it should be noted that the original research developing the MOXFQ utilised a cohort of foot surgery patients with post- operative follow-up set at 12 months.

An initial scoping literature search by the lead author highlighted 44 articles concerned with the outcomes of foot surgery within a two-year period. Of 23 unique tools, the top three were the American Orthopaedic Foot & Ankle Society (AOFAS) (10), the Visual Analogue Scale (VAS) (11) and the Manchester-Oxford Foot/Ankle Questionnaire (MOXFQ – see Appendix 1) (12), which together account for 66% usage of all the tools used. In 2018, a systematic review of five years of data by Shazadeh Safavi et al. (13) reported on 76 tools; in 2019, the review by Lakey & Hunt (14) reported on 89 tools.

AOFAS has not been demonstrated to be either valid or reliable, and in 2011 the AOFAS organisation released a statement recommending discontinuing the use of the system (15). The VAS is a generic instrument, non-specific to a disease or foot region, with low specificity for minimally clinically important differences (MCID) (14). The MOXFQ, by comparison, has been tested for validity, reliability, and responsiveness in the context of foot surgery (16). Its use has increased in popularity from 1% use in the review by Hasenstein et al. (7) in 2017 to 39% in the senior author’s initial search. The MOXFQ questionnaire has been tested and validated for use with all foot surgery (16). There are 16 questions using a Likert scale, with responses assigned to one of three domains: walking/standing (WS), pain (P) and social interaction (SI).

Patients’ health-related quality of life (HRQOL) is liable to change with time, although data from studies are conflicting. Though not directly relevant, the retrospective study of patient satisfaction by Taylor et al. (9) using the PASCOM-10 patient satisfaction questionnaire (PSQ-10) with a follow-up time ranging from seven to 82 months found no statistically significant change to satisfaction scores at differing time points. However, the authors advised felt six months was adequate.

This study aimed to determine if the patient’s health-related quality of life as measured by the MOXFQ changes after foot surgery with differing time intervals in a cohort of podiatric surgery patients. The hypothesis this study sets out is that there will be no differences in patient health-related quality of life after foot surgery with differing time intervals.

## Method

The study is a retrospective, quantitative audit to assess patient health-related quality of life before surgery and at six and 12 months in one NHS podiatric surgery centre (Northamptonshire) in the UK. Two hundred and sixty-nine participants were identified (convenience sample) from a retrospective screen of the PASCOM-10 surgical database from January to December 2019. The authors considered that the 12-month period would generate an acceptable cohort of participants. The participants were then cross-checked to ensure that pre-operative MOXFQ values had been recorded. Following this, 33 blank forms, duplicate entries and data errors were excluded. A total of 236 patients were included in the cohort; see flow chart in Figure 1.

**Figure 1:**
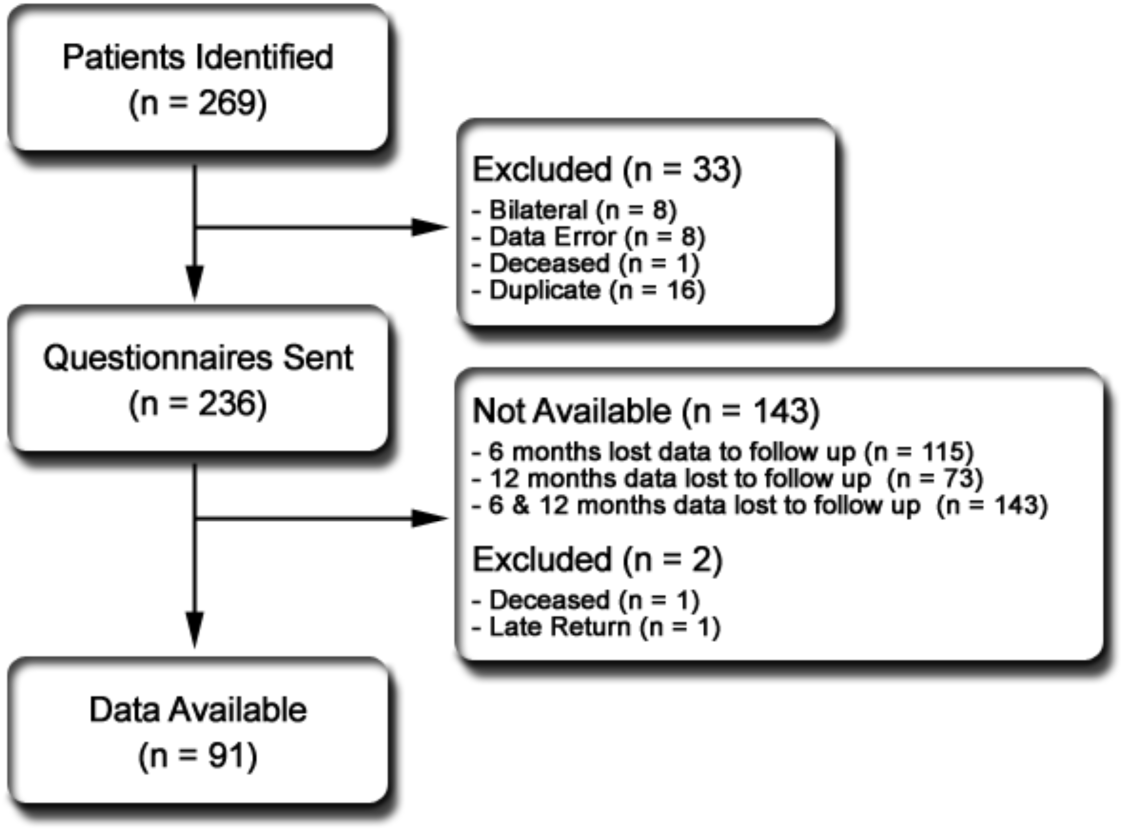
participant flowchart.

Two hundred thirty-six questionnaires were prepared for the 12-month point for all participants; these questionnaires were sent through the post with free postage returns at the appropriate 12-month post-surgery interval. The following information was extracted from PASCOM-10 and exported to a Microsoft Excel spreadsheet for further analysis: gender, age, surgical complications, type of foot surgery, anatomic location, and MOXFQ pre and six-month values.

Table 1 shows the 16 MOXFQ questions segregated by the three domains of walking/standing (WS), pain (P) and social interaction (SI) and the associated Minimum Clinically Important Difference (MCID). That is the threshold score at which the patient notices and actual rather than statistical improvement in HRQOL. The allocation of questions to each domain means they have differing totals; percentages are calculated and used for comparison.

**Table 1:** MOXFQ Question numbers, categories and MCID (12)

| Question Numbers | Category | MCID |
| --- | --- | --- |
| 2, 3, 4, 5, 6, 7, 8 | walking/standing | 16 |
| 1, 11, 12, 15, 16 | pain | 12 |
| 9, 10, 13, 14 | social interaction | 24 |

Data analysis was conducted in Microsoft Excel and Access, and all descriptive statistics using SPSS version 26.0 (IBM Corp) following the parsimonious model, significance level .05, confidence interval 95%. Analysis of variance (ANOVA) was used with the Bonferroni adjustment. The Huynh-Feldt correction was used where the Mauchly test of sphericity was violated, and where an interaction effect was found, an analysis of simple effects was performed. Tests of within subjects’ effects used the Wilks Lambda test.

Repeated measures mixed analyses of variance were performed on all 91 participants with data from all time points to analyse if there were any significant changes in time or differences in outcome between the treatment groups for domains of WS, P and SI, age, gender, or treatment type, location, and sequelae over time. The variables deemed to be of interest for this research included the MOXFQ domains of WS, P and SI alongside gender and all time points (0, 6, and 12 months).

## Results

163 questionnaires were returned (61%), one deceased, one late return and 73 lost to follow-up. The 6-month data were collected during the post-operative appointment. These appointments were not available after April 2020 due to Corona virus restrictions and thus were sent out through the post. This affected 76 participants. 120 questionnaires were completed (45%), 115 lost to follow-up. 91 participants (39%) completed both six-month and 12-month questionnaires (66 female and 25 male. The mean age is 64, range 35-93 years). The anatomical location of the surgical procedure on the foot was too small to be analysed individually and was grouped (forefoot hallux, forefoot and midfoot/rearfoot). Surgical procedures and post- operative complications had insufficient numbers to enable statistical analysis (16). There were 76 procedure variations. Table 2 summarises the mean descriptive statistics for the MOXFQ at all time points and genders.

**Table 2:**
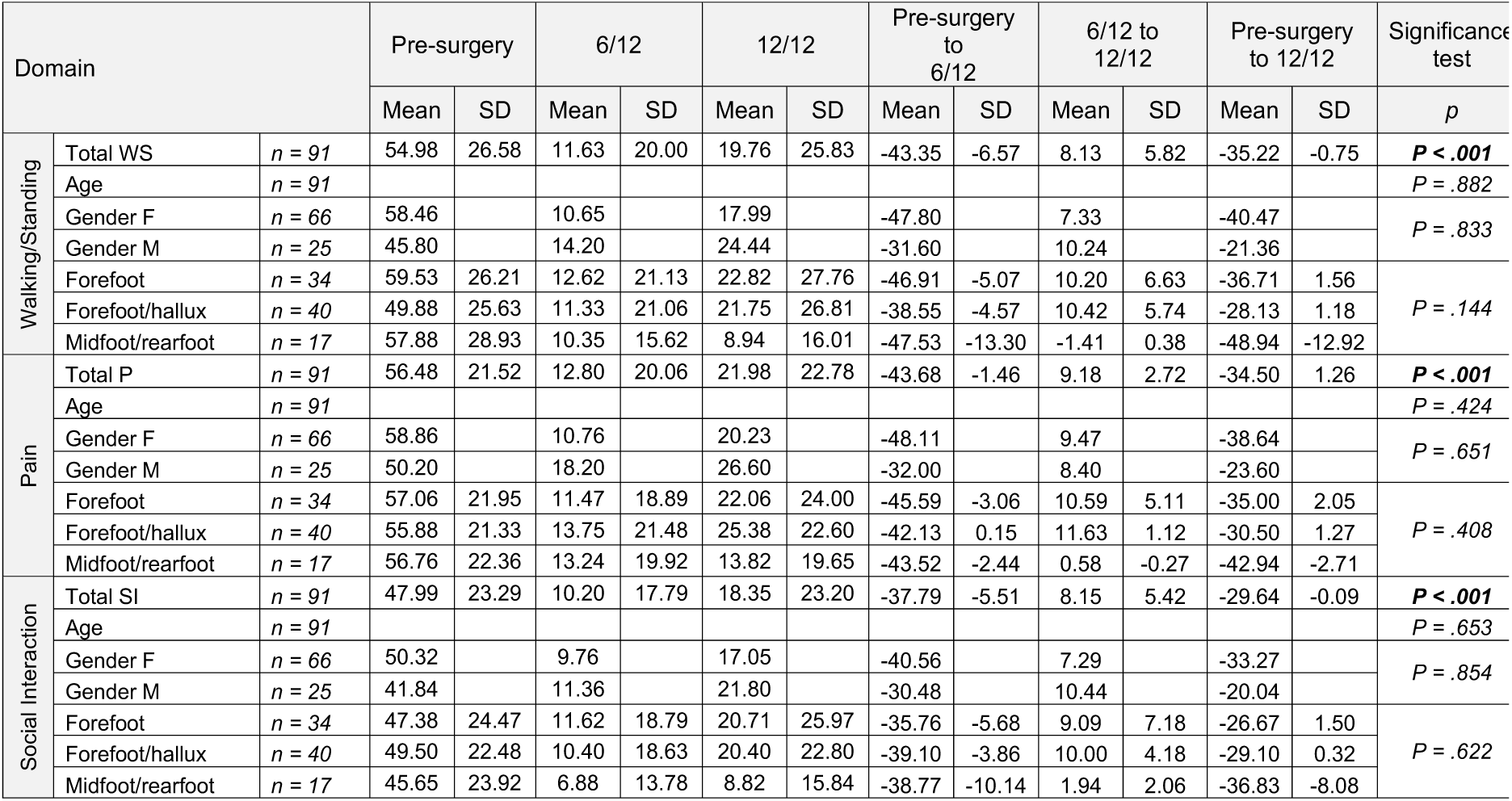
mean descriptive statistics for the MOXFQ.

Mauchly’s test indicated that the assumption of sphericity had been violated in all findings, and the degrees of freedom were subsequently corrected using Huynh-Feldt estimates of sphericity, results are given via a variable for the three MOXFQ domains of WS, P and SI.

There was no correlation between the age of the participant and the values at different time points, F(1.69,149.99) = .092, p = .882, F(1.74,154.5) = .827, p = .424, F(1.58,136.77) = .351, p = .653 WS, P and SI respectively. The results show that there were no significant effect of anatomic location of surgery with any time point and the three MOXFQ domains, F(3.42,150.42) = 1.79, p = .144, F(3.50,153.79) = .99, p = .41, F(3.17,139.57) = .61, p = .622 WS, P and SI respectively. Age and anatomic location had no effect on MOXFQ scores.

A significant interaction was found with all three MOXFQ domains over all the time points (pre-surgery, six- and 12 months), and a correlation was found. Wilks’ Lambda = 0.353, F(6, 356) = 40.59, p <.001, and for all time points in combination, shown in Figure 2, Figure 3 and Table 3.

**Figure 2.**
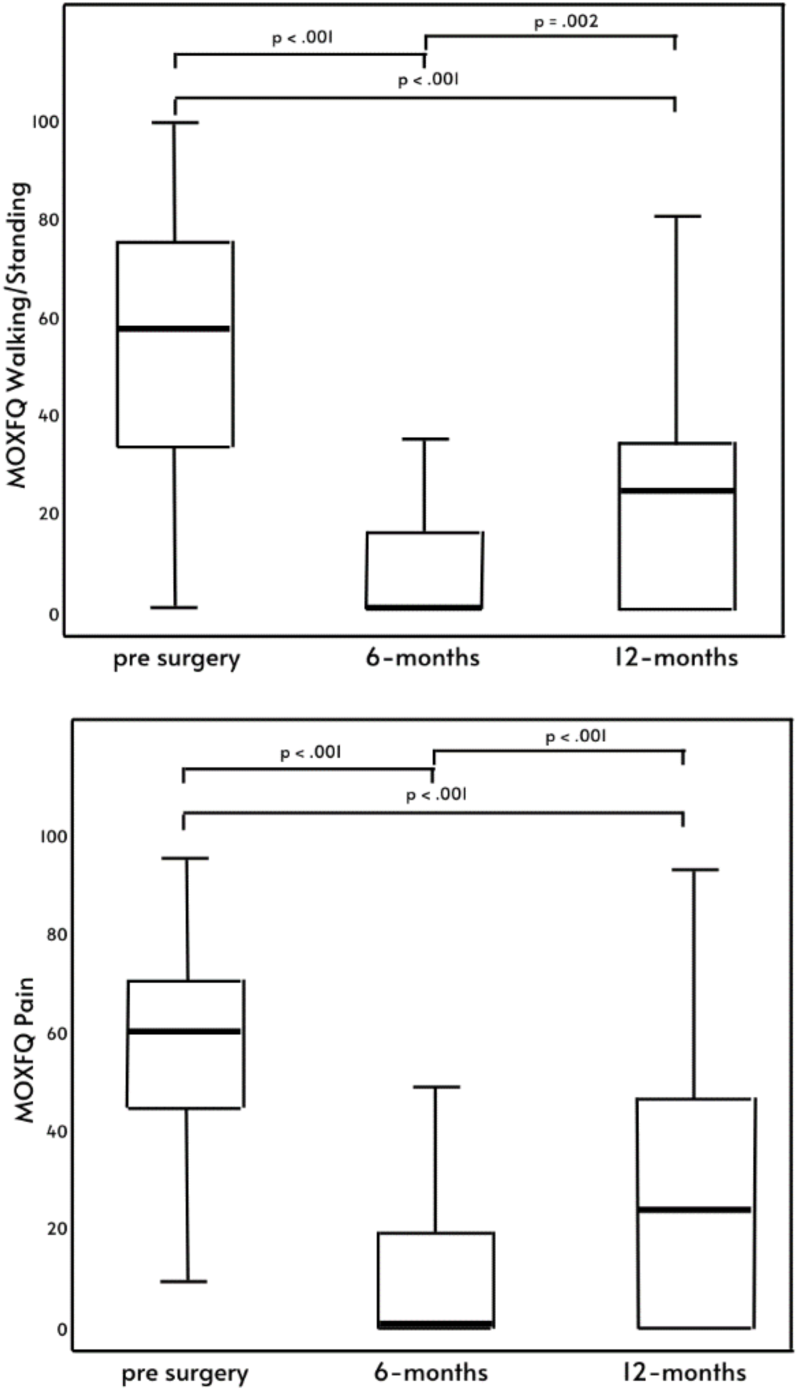
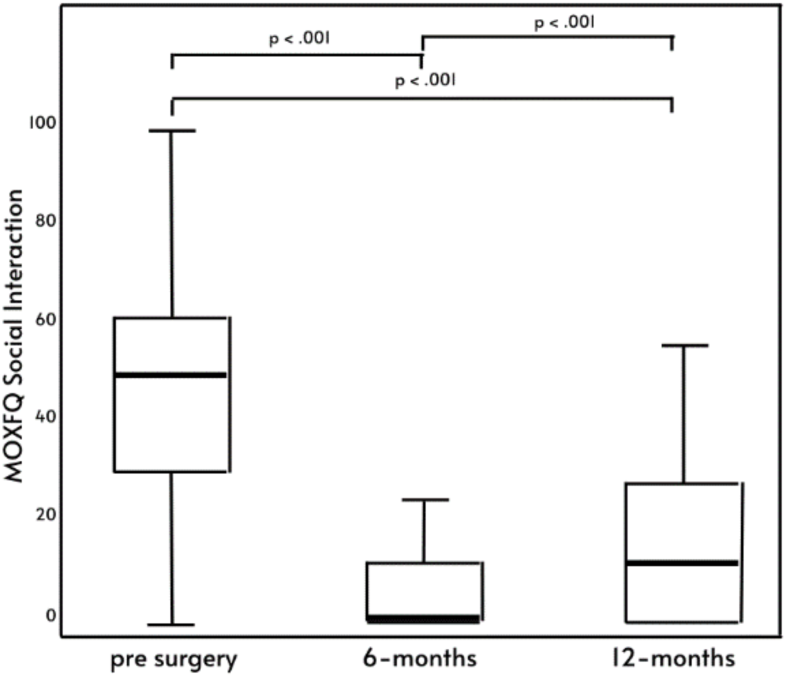
(a, b, c): Domains p-values at time points created using SPSS.

**Figure 3:**
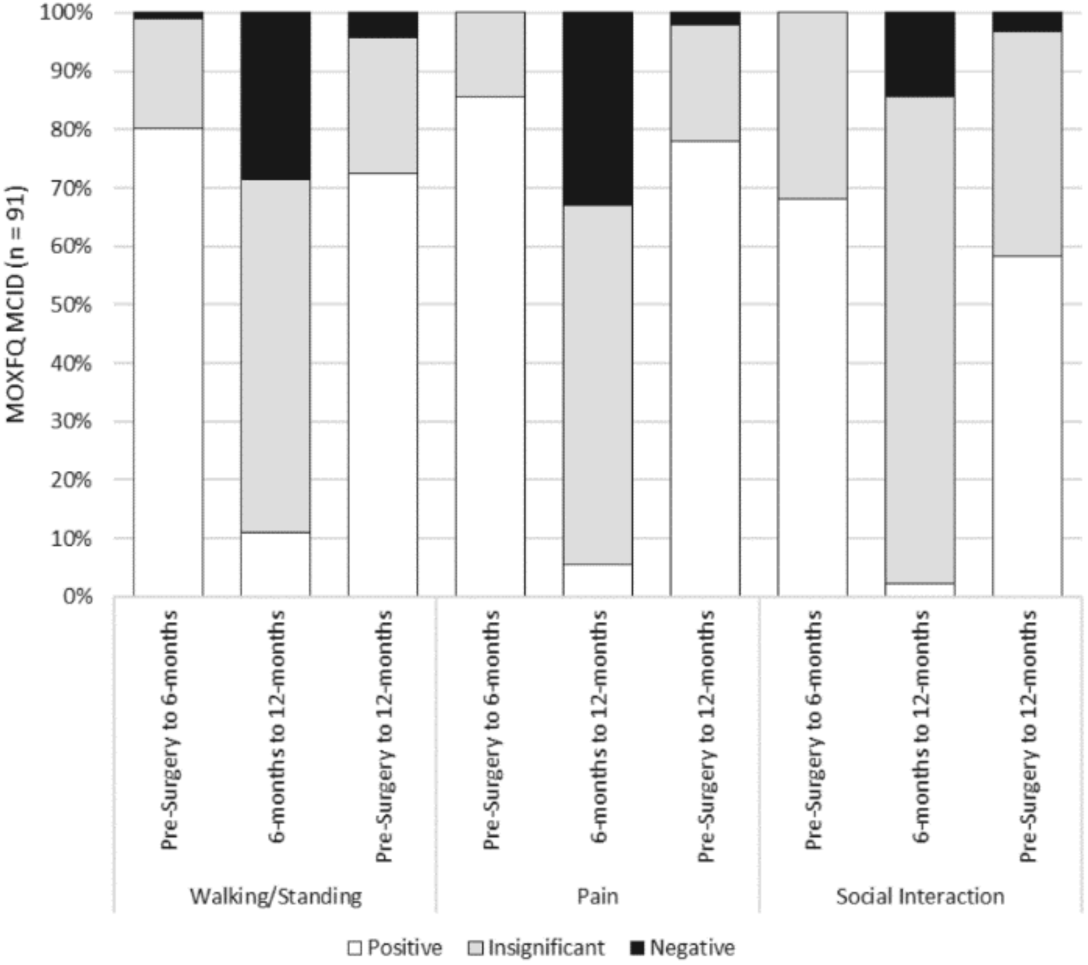
MCID change in domains at time points.

**Table 3:** Breakdown of the analysis of MOXFQ scores across the domains.

| *n* = 91 | *Multivariate* | Univariate |  |  |
| --- | --- | --- | --- | --- |
|  |  | Walking/Standing | Pain | Social Interaction |
| Time Points | $F(6, 356) = 40.59$, **$P < .001$** | $F(1.67, 149.90) = 115.15$, **$P < .001$** | $F(1.72, 155.2) = 157.5$, **$P < .001$** | $F(1.59, 140.22) = 117.93$, **$P < .001$** |
| Gender | $F(6, 352) = 30.23$, **$P < .001$** | $F(1.72, 153.27) = 4.773$, **$P = .013$** | $F(1.78, 158.54) = 5.01$, **$P = .010$** | $F(1.6, 142) = 2.898$, $P = .070$ |
| Age | $F(6, 352) = .574$, $P = .75$ | $F(1.69, 149.99) = .092$, $P = .882$ | $F(1.74, 154.51) = .827$, $P = .424$ | $F(1.58, 140.26) = .351$, $P = .653$ |
| Location | $F(12, 460.65) = .111$, $P = .353$ | $F(3.42, 150.42) = 1.79$, $P = .144$ | $F(3.50, 153.79) = .99$, $P = .41$ | $F(3.17, 139.57) = .61$, $P = .622$ |

Multivariate results reported using Wilks Lambda. **Bold is significant to $P < .05$ .**

There was a significant interaction between gender and time points for domains WS and P, F(1.72,153.27) = 4.773, p = .013, F(1.78,158.54) = 5.01, p = .010 (Table 4), however there was no significant effect with SI and time points, WS, P and SI (P=0.833, .651, .854 respectively). The interaction of time and gender is non-significant, Wilks’ Lambda = 0.935, F (6,352) = 2.02, p = 0.62, although there was a significant effect of gender with linear effect at all time points, Wilks’ Lambda = 0.435, F (6,352) = 30.23, p <.001. Time is shown in Figures 4 and 5.

**Figure 4:**
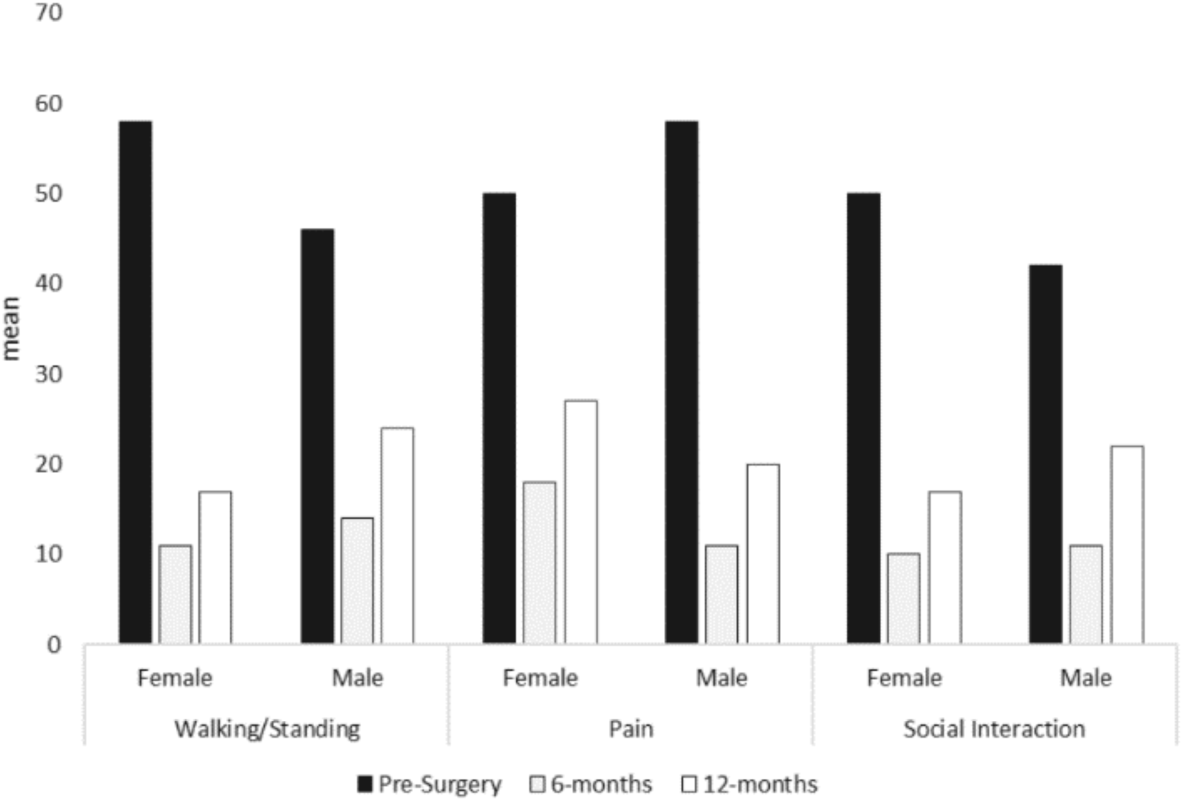
MOXFQ domain mean scores split by gender.

**Figure 5.**
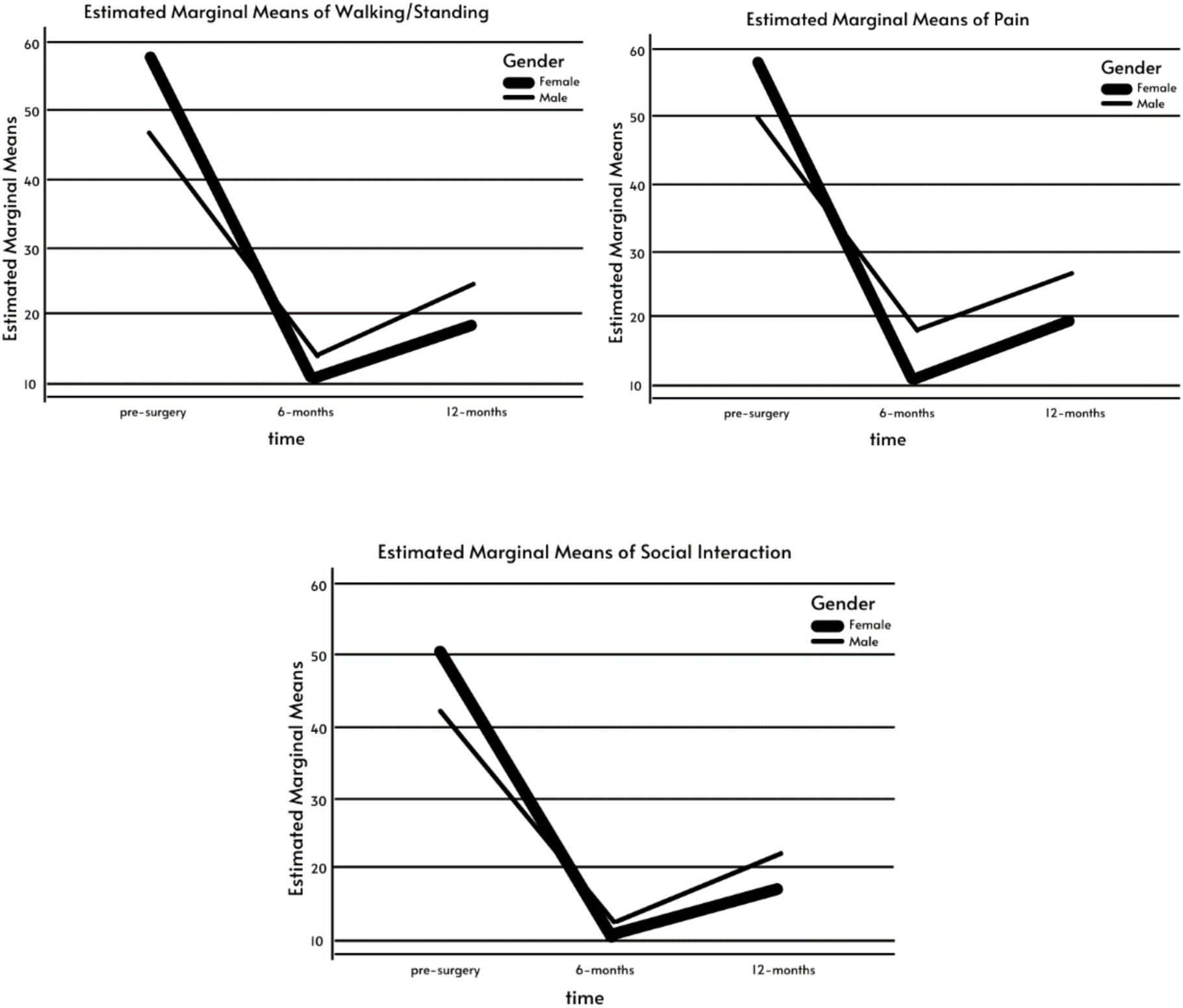
(a, b, c): MOXFQ domain scores by gender at time points.

**Table 4:** p-value of the analysis of MOXFQ scores across the three domains.

|  | Between subject effects |  |  |
| --- | --- | --- | --- |
|  | Walking/Standing | Pain | Social Interaction |
| Pre-surgery to 6/12 | ***P* < .001** | ***P* < .001** | ***P* < .001** |
| 6/12 – 12/12 | ***P* = .002** | ***P* < .001** | ***P* < .001** |
| Pre-surgery to 12/12 | ***P* < .001** | ***P* < .001** | ***P* < .001** |
| Gender | *P* = .833 | *P* = .651 | *P* = .854 |

**Table 5:**
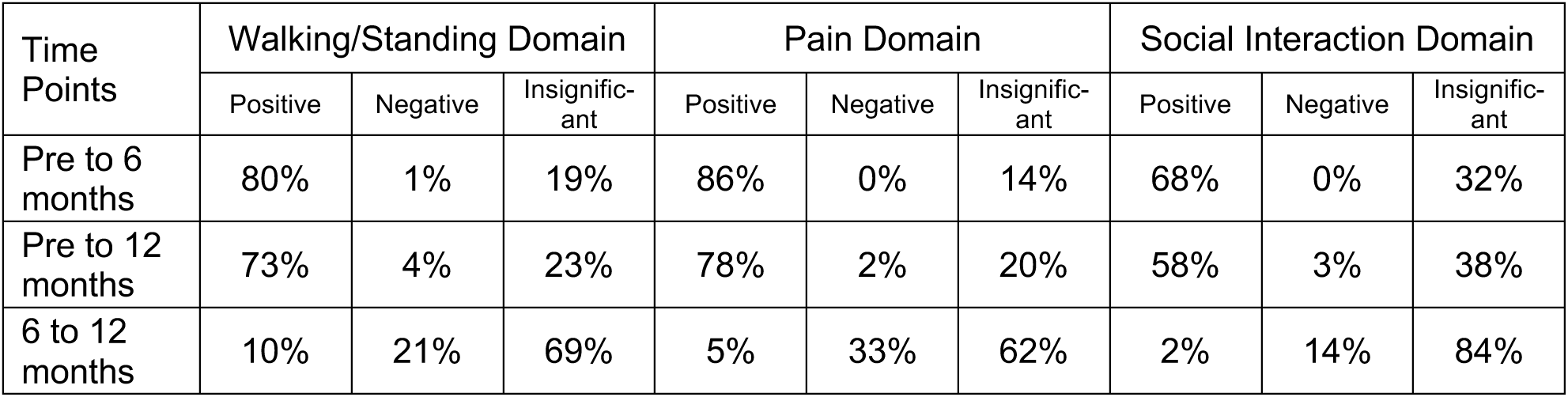
MCID Percentage Change in domain scores at all time points.

| Time Points | Walking/Standing Domain |  |  | Pain Domain |  |  | Social Interaction Domain |  |  |
| --- | --- | --- | --- | --- | --- | --- | --- | --- | --- |
|  | Positive | Negative | Insignific-ant | Positive | Negative | Insignific-ant | Positive | Negative | Insignific-ant |
| Pre to 6 months | 80% | 1% | 19% | 86% | 0% | 14% | 68% | 0% | 32% |
| Pre to 12 months | 73% | 4% | 23% | 78% | 2% | 20% | 58% | 3% | 38% |
| 6 to 12 months | 10% | 21% | 69% | 5% | 33% | 62% | 2% | 14% | 84% |

Adjustment for multiple comparisons: Bonferroni. Based on estimated marginal means - the mean difference is significant at the .05 level.

Female HRQOL appeared poorer than males across all three domains at pre-surgery and at 12 months. Female HRQOL from pre-surgery to 6 months shows the greatest gain (p=.013), 82%, 82% and 81% compared to males 69%, 64% and 73%, for WS, P and SI. From six to 12 months both genders see an increase in MOXFQ scores across all three domains, equating to a reduction in HRQOL (p=0.10).

## Discussion

The three MOXFQ domains show variations at different time points, all with statistical significance, and all follow the same trajectory. At six months post podiatric surgery, patients experience the greatest positive change in HRQOL scores, an average improvement of 78% across all domains. Health-related quality of life deteriorates during the time period of six to 12 months after surgery, and at 12 months the HRQOL score is an average of 62% improved over the preoperative scores.

The need for assessment of patient-reported outcomes at different time intervals has been suggested by The Health Foundation (2) which recommends a need for further research in assessing the quality of health services over time. 12 months is recommended by Dawson, Coffey et al in their original study looking at MOXFQ responsiveness and the use of MCID (16). In contrast, the study by Taylor et al. (9) using only the PASCOM PSQ-10 questionnaire reported positive changes over time (beyond six months) but concluded that six months is an adequate period to analyse patient progress and likely representative of the final outcome. Due to the retrospective nature of data collection, their study was unable to specify exact follow- up time points, and the sensitivity of the data is unknown.

The MOXFQ employs three separate domains relevant to foot health to explore HRQOL, all three domains at all three time points have been found to have significant findings. Of interest, the improvement in HRQOL appears to peak at six months before reducing slightly at 12 months. The score changes across the three domains between six and 12 months were significant however, that reduced 12-month MOXFQ score still represents an improvement in HRQOL over the pre-operative state.

From surgery to six months post-operation patients are given guidelines to follow including activity modification, footwear advice, wound care, and pain management. At six months post-operation patients are typically discharged and informed to start returning to normal activity. As patients return to their pre-surgery activities this may contribute to foot pain as the foot is still arguably healing. That pain may in turn limit participation in weight-bearing activities or at least make those activities more difficult which may in turn account for the reduction in domain scores. High-impact and contact sports in particular report the most negative outcomes following treatment (17).

An interesting finding of the present study was the observed difference in HRQOL scores between males and females. At the time of surgery, females had significantly poorer scores across all three MOXFQ domains than their male counterparts. Females then went onto see more significant improvement in HRQOL at 6 months before deteriorating slightly at 12 months. The study by MacInnes and Roberts (17) noted a non-significant mean difference between genders of only 17%, unlike 30% in this study.

A number of studies recommend a follow-up longer than six months, as certain procedures may require longer periods to fully rehabilitate, such as ankle fusions and getting back into sports (5). Maher and Kilmartin (8) warn that assessing patients early may not provide a realistic gauge of the ultimate outcome and that for reconstructive foot surgery patients will continue to improve after 12 months. Taylor et al. (9) discuss forefoot surgery requiring six months, midfoot and hindfoot six-18 months discharge. This study was unable to conclude on surgery type for the results and would benefit from further study in this area.

The experimenter effect can result in subtle differences in participant treatment. At the six months point, the surgeon would discuss the outcomes and his thoughts before requesting the completion of the questionnaire, thus possibly influencing the participants scoring. The 12-month point questionnaires were completed through the post. The six month scores may be a direct consequence of the experimenter effect.

The six month questionnaires have a lower completion number than the 12 month data and after April 2020 when the unit closed due to the COVID-19 pandemic. The average 6-month completion of questionnaires for 2019 was 55%, including 63% before closing dropping to 38% after closure. It is unknown how the missing data may have affected the results.

There may be a responder bias from the use of postal questionnaires, with the additional disadvantage of a low response rate and patients not included owing to missing follow-up data, a change of address or lack of interest. The authors believe this is in part due to the study being performed at the height of the COVID-19 pandemic.

## Conclusion

The results of this study demonstrate that at different time points, patients report different MOXFQ scores. A significant improvement in MOXFQ scores across all three domains is noted from pre-surgery to six months, with a reduction by the time 12 months have elapsed. The most significant improvement is seen in females compared to males, with a 30% difference. However, females start and finish with lower health-related quality of life than men.

The current six-month point for PROMs to be recorded by the surgical team may not provide the most accurate result for the patient or clinician. Post-operative recovery is not simply a period when all wounds heal, and all pain dissipates; it is a period that may last longer than 6 months. Patients need to be aware that six months may not provide the optimum recovery, and once the 6-month stage has been reached, it may take time to settle. Whilst acknowledging that different data collection methods may have affected scores, this study recommends extending the six-month data collection point for the MOXFQ questionnaires to twelve months.

## Data Availability

Availability of data and material: anonymised data on PASCOM-10.

## Acknowledgements

The authors would like to thank John Stephenson (University of Huddersfield) for his assistance with statistical analysis.

## Author information

Northamptonshire Healthcare Foundation NHS Trust, Danetre Hospital, Daventry, Northamptonshire. NN11 4DY. UK VN: https://orcid.org/0000-0002-0835-866X INR: https://orcid.org/0000-0002-2786-5739 AB: https://orcid.org/0000-0002-4942-7051 AM: https://orcid.org/0000-0002-9173-2981

## List of abbreviations

AOFAS: American Orthopaedic Foot & Ankle Society
CQC: Care Quality Commission
FFI: Foot Function Index
HRQOL: Health-related quality of life
MOXFQ: Manchester Oxford foot questionnaire
MCID: Minimally clinically important difference
PROMs: Patient reported outcomes measures
PASCOM-10: Podiatric Audit of Surgery and Clinical Outcome Measurement
PSQ-10: Patient satisfaction questionnaire
VAS/VAS FA: Visual analogue scale/foot and ankle

## Declarations

### Ethics approval and consent to participate

Ethical approval for this study was granted on 14.02.20 from the NHFT Research and Innovation Department in compliance with NHFT’s Policy for Conducting Clinical Audit Projects (CLP019). Further information/documentation is available on request.

Consent for publication was granted via NHFT Research and Innovation Department.

### Availability of data and material

Availability of data and material: anonymised data on PASCOM-10.

### Authors’ contributions

Author information: IR conceived the aim and format of the paper. VN performed the literature search and produced the first draft. All authors made substantial contributions to the final version.

---

**Appendix 1:** Manchester Oxford Foot Questionnaire.

| Circle as appropriate:<br>RIGHT / LEFT FOOT<sup>1</sup><br>During the past 4 weeks this<br>has applied to me: | Please tick ✓ one box for each statement |  |  |  |  |
| --- | --- | --- | --- | --- | --- |
|  | None of<br>the time | Rarely | Some of<br>the time | Most of<br>the time | All of<br>the time |
| 1. I have pain in my foot | ☐ | ☐ | ☐ | ☐ | ☐ |
| 2. I avoid walking long distances because of pain in my foot | ☐ | ☐ | ☐ | ☐ | ☐ |
| 3. I change the way I walk due to pain in my foot | ☐ | ☐ | ☐ | ☐ | ☐ |
| 4. I walk slowly because of pain in my foot | ☐ | ☐ | ☐ | ☐ | ☐ |
| 5. I have to stop and rest my foot because of pain | ☐ | ☐ | ☐ | ☐ | ☐ |
| 6. I avoid some hard or rough surfaces because of pain in my foot | ☐ | ☐ | ☐ | ☐ | ☐ |
| 7. I avoid standing for a long time because of pain in my foot | ☐ | ☐ | ☐ | ☐ | ☐ |
| 8. I catch the bus or use the car instead of walking, because of pain in my foot | ☐ | ☐ | ☐ | ☐ | ☐ |
| 9. I feel self-conscious about my foot | ☐ | ☐ | ☐ | ☐ | ☐ |
| 10. I feel self-conscious about the shoes I have to wear | ☐ | ☐ | ☐ | ☐ | ☐ |
| 11. The pain in my foot is more painful in the evening | ☐ | ☐ | ☐ | ☐ | ☐ |
| 12. I get shooting pains in my foot | ☐ | ☐ | ☐ | ☐ | ☐ |
| 13. The pain in my foot prevents me from carrying out my work/everyday activities | ☐ | ☐ | ☐ | ☐ | ☐ |
| 14. I am unable to do all my social or recreational activities because of pain in my foot | ☐ | ☐ | ☐ | ☐ | ☐ |
| 15. During the past 4 weeks how would you describe the pain you usually have in your foot? (please tick one box) | None<br>☐ | Very mild<br>☐ | Mild<br>☐ | Moderate<br>☐ | Severe<br>☐ |
| 16. During the past 4 weeks have you been troubled by pain from your foot in bed at night? (please tick one box) | No nights<br>☐ | Only 1 or 2 nights<br>☐ | Some nights<br>☐ | Most nights<br>☐ | Every night<br>☐ |

<sup>1</sup>The foot to be assessed may be indicated here. Alternatively, each question may be customised to the right foot with all questions then repeated and customised to the left foot.
Finally, please check that you have answered every question
Thank you very much

## References

1. Maher AJ, Metcalfe SA. A report of UK experience in 917 cases of day care foot surgery using a validated outcome tool. The Foot. 2009;19:6. https://doi.org/10.1016/j.foot.2009.01.002.

2. Appleby J. Day case surgery: a good news story for the NHS. BMJ: British Medical Journal. 2015 Jul 29;351:h4060. https://www.jstor.org/stable/26522549.

3. The King’s Fund. The NHS in a nutshell. 2020. Available from: https://www.kingsfund.org.uk/projects/nhs-in-a-nutshell/NHS-activity Accessed 05.04.22.

4. Chow A, Mayer EK, Darzi AW, Athanasiou T. Patient-reported outcome measures: the importance of patient satisfaction in surgery. Surgery. 2009 Sep 1;146(3):435–43. https://doi.org/10.1016/j.surg.2009.03.019.

5. Dawson J, Boller I, Doll H, Lavis G, Sharp RJ, Cooke P, Jenkinson C. Factors associated with patient satisfaction with foot and ankle surgery in a large prospective study. The Foot. 2012 Sep 1;22(3):211–8. https://doi.org/10.1016/j.foot.2012.05.002.

6. Maher A. Service Evaluation, outcome measurement and PASCOM-10. Podiatry Now. 2016;December (12):16–19.

7. Hasenstein T, Greene T, Meyr AJ. A 5-Year review of clinical outcome measures published in the Journal of the American Podiatric Medical Association and the Journal of Foot and Ankle Surgery. The Journal of Foot and Ankle Surgery. 2017 May 1;56(3):519–21. https://doi.org/10.7547/16-157.

8. Maher AJ, Kilmartin TE. An analysis of Euroqol EQ-5D and Manchester Oxford Foot Questionnaire scores six months following podiatric surgery. Journal of Foot and Ankle Research. 2012;5(1):1–7. https://doi.org/10.1186/1757-1146-5-17

9. Taylor NG, Tollafield DR, Rees S. Does patient satisfaction with foot surgery change over time? The Foot. 2008;18(2):68–74. https://doi.org/10.1016/j.foot.2008.01.003.

10. Kitaoka HB, Alexander IJ, Adelaar RS, Nunley JA, Myerson MS, Sanders M. Clinical rating systems for the ankle-hindfoot, midfoot, hallux, and lesser toes. Foot & Ankle International. 1994 Jul;15(7):349–53. https://doi.org/10.1177/1071100794015007.

11. Hayes MH, Patterson DG. Experimental development of the graphic rating method. Psychological Bulletin. 1921,18:98–99.

12. Dawson J, Boller I, Doll H, Lavis G, Sharp R, Cooke P, Jenkinson C. The MOXFQ patient-reported questionnaire: Assessment of data quality, reliability and validity in relation to foot and ankle surgery, The Foot, 21(2):92–102, https://doi.org/10.1016/j.foot.2011.02.002.

13. Shazadeh Safavi P, Janney C, Jupiter D, Kunzler D, Bui R, Panchbhavi VK. A systematic review of the outcome evaluation tools for the foot and ankle. Foot & Ankle Specialist. 2019 Oct;12(5):461–70. https://doi.org/10.1177/1938640018803747.

14. Lakey E, Hunt KJ. Patient-reported outcomes in foot and ankle orthopedics. Foot & Ankle Orthopaedics. 2019 Jul 12;4(3):2473011419852930. https://doi.org/10.1177/2473011419852930.

15. Pinsker E, Daniels TR. AOFAS position statement regarding the future of the AOFAS Clinical Rating Systems. Foot & Ankle International. 2011 Sep;32(9):841–2. https://doi.org/10.3113/FAI.2011.0841.

16. Dawson J, Doll H, Coffey J, Jenkinson C. Responsiveness and minimally important change for the Manchester-Oxford foot questionnaire (MOXFQ) compared with AOFAS and SF-36 assessments following surgery for hallux valgus. Osteoarthritis and Cartilage. 2007 Aug 1;15(8):918–31. https://doi.org/10.1016/j.joca.2007.02.003

17. MacInnes A, Roberts SC, Kimpton J, Pillai A. Long-term outcome of open plantar fascia release. Foot & Ankle International. 2016 Jan;37(1):17–23. https://doi.org/10.1177/1071100715603189.

